# No Evidence of Infectious SARS-CoV-2 in Human Milk: Analysis of a Cohort of 110 Lactating Women

**DOI:** 10.1101/2021.04.05.21254897

**Authors:** Paul Krogstad, Deisy Contreras, Hwee Ng, Nicole Tobin, Christina D. Chambers, Kerri Bertrand, Lars Bode, Grace Aldrovandi

## Abstract

**Background:** SARS-CoV-2 infections of infants and toddlers are usually mild but can result in life-threatening disease. SARS-CoV-2 RNA been detected in the breast milk of lactating women, but the potential role of breastfeeding in transmission to infants has remained uncertain.

**Methods:** Breast milk specimens were examined for the presence of the virus by RT-PCR and/or culture. Specimens that contained viral RNA (vRNA) were examined for the presence of subgenomic coronavirus RNA (sgRNA), a putative marker of infectivity. Culture methods were used to determine the thermal stability of SARS-CoV-2 in human milk.

**Results:** Breast milk samples from 110 women (65 confirmed with a SARS-CoV-2 diagnostic test, 36 with symptoms but without tests, and 9 with symptoms but a negative SARS-CoV-2 diagnostic test) were tested by RT-PCR (285 samples) and/or viral culture (160 samples). Although vRNA of SARS-CoV-2 was detected in the milk of 7 of 110 (6%) women with either a confirmed infection or symptomatic illness, and in 6 of 65 (9%) of women with a positive SARS-CoV-2 diagnostic test, virus was not detected in any culture. None of the 7 milk specimens with detectable vRNA contained sgRNA. Notably, when artificially added to human milk in control experiments, infectious SARS-CoV-2 could be cultured despite several freeze-thaw cycles, as occurs in the storage and usage of human milk.

**Conclusions:** SARS-CoV-2 RNA can be found infrequently in the breastmilk of women with recent infection, but we found no evidence that breastmilk contains infectious virus or that breastfeeding represents a risk factor for transmission of infection to infants.

**Key Points:** *Question:* SARS-CoV-2 RNA has been detected in a small number of human milk samples collected from recently infected women. The role of breastfeeding in transmission of the virus to infants has remained uncertain due to the small number of specimens analyzed in any study published thus far.

*Findings:* In a total study group of 110 women, SARS-CoV-2 RNA was detected in milk from 6 of 65 women (9.2%) with recent confirmed infection. Neither infectious virus nor subgenomic RNA (a marker of virus infectivity) were detected in any of the samples.

*Meaning:* We found no evidence that infectious SARS-CoV-2 is present milk from recently infected women, even if SARS-CoV-2 PCR tests are positive, providing reassurance of the safety of breastfeeding.

## INTRODUCTION

To date, the COVID-19 pandemic has infected nearly 100 million people globally and over 21 million people in the United States ^1^. COVID-19 cases in infants represent about 50% of the total reported in young children globally ^2^, and infections of infants represent approximately 1.9% of all cases in the United States ^3^. Most newborns and infants fare well but severe disease and death have been reported in newborns, infants and young children ^2^. In addition, multisystem inflammatory syndrome in children (MIS-C) can occur even after apparent resolution of infection and is disproportionately affecting Black and Hispanic/Latino children ^4^. Understandably, there has been great concern about the potential consequences of SARS-CoV-2 transmission to infants by postnatal exposures, including breast feeding. There is limited but increasing evidence that breast milk is not a source of SARS-CoV-2 infection to infants who are breastfed by mothers who have evidence of recent infection ^5,6^. Therefore, the U.S. Centers for Disease Control and Prevention, the American Academy of Pediatrics and the World Health Organization advise that mothers who are infected with SARS-CoV-2 may choose to initiate or to continue breastfeeding an infant or toddler with appropriate protections to prevent transmission of the virus through respiratory droplets^7,8^.

We previously presented initial results from a study of 64 breast milk samples from 18 women with recent SARS-CoV-2 infection ^9^. A sample from one symptomatic woman was found to contain SARS-CoV-2 viral RNA, but replication competent virus was not detected in viral culture. In addition, we used viral culture methods to demonstrate that the conditions of Holder pasteurization inactivated SARS-CoV-2, a finding subsequently confirmed by others ^10-12^. To better examine the frequency and state of SARS-CoV-2 in breast milk of women with recently documented infection, we present results from a larger observational cohort study. We also examined the viral RNA found in RT-PCR positive samples for the presence of SARS-CoV-2 subgenomic transcripts, a proposed marker of viral infectivity ^13-15^. In addition, we examined the impact of breast milk on SARS-CoV-2 thermal stability to investigate the apparent discordance between viral culture and viral RNA detection.

## MATERIALS AND METHODS

### Participants and Breast Milk Specimens

As previously described ^9^ breast milk samples and clinical information were obtained from women participating in the Mommy’s Milk Human Milk Biorepository at the University of California, San Diego (IRB#130658). Women residing in the United States who were symptomatic but not tested, symptomatic but with negative SARS-CoV-2 testing by RT-PCR, exposed to an infected person, or those who had a confirmed SARS-CoV-2 infection by RT-PCR were enrolled into the study between March 2020 and September 2020. Information about demographics, health history, illness and exposure dates, symptoms and SARS-CoV-2 test results were collected by participant interview via telephone. Participants self-collected breast milk samples using a provided collection kit including instructions for expressing and storing their samples. Instructions included hand washing before and after milk expression. Participants who had recovered from their illness at the time of the study interview were asked to ship any frozen samples previously collected at the peak of their symptoms in addition to a fresh milk sample. Fresh samples were shipped on ice within 24h of collection to the Biorepository and stored at ^-^80°C prior to shipment on dry ice to the University of California, Los Angeles. This report includes samples from 18 women described in our previous report^9^.

### Virologic Methods

The molecular methods used to detect SARS-CoV-2 RNA in skim milk and the culture techniques to detect replication competent SARS-COV-2 in whole breast milk have been previously described ^9^. The concentration of replication competent virus in SARS-COV-2 viral stocks was determined by limiting dilution culture, calculated using the Spearman-Karber method, and expressed as the median tissue culture infectious doses (TCID_50_), as before. Detection of subgenomic SARS-CoV-2 RNA (a proposed marker of viral replication ^13-15^) was performed using oligonucleotide PCR primers WHSA-00025F and WHSA29925R ^13^ and a Power Syber green RNA to Ct 1-step kit (Thermo Fisher Scientific), following the manufacturer’s instructions.

### Thermal Stability of SARS-CoV-2

We previously reported that SARS-CoV-2 could not be grown from a whole breast milk specimen that was shown to contain a high concentration of SARS-CoV-2 RNA^9^. In that report we also noted that the temperature conditions of Holder Pasteurization rapidly inactivated live virus, even if diluted ten-fold in breast milk from healthy donors. However, it remained possible that we were unable to grow SARS-CoV-2 due to the presence of antimicrobial factors in breast milk that could reduce viral infectivity during the cycles of freezing and gradual low temperature thawing that commonly occur with storage and preparation of breast milk. To examine the thermal stability of the virus during these steps, we added a small amount (100 TCID_50_) of SARS-CoV-2 (USA-WA1/2020) to 4 aliquots each of whole breast milk from two different healthy women. One aliquot from each set was then held at 4°C, and three others were frozen again at ^-^80°C. These aliquots were slowly thawed to room temperature (approximately 20°C). Two aliquots were frozen again to ^-^80°C, and one was held at 4°C. This process was repeated, yielding samples that had undergone 2 freeze-thaw cycles. Following a third freeze and thaw for aliquots from both women, all samples were stored at 4°C for 3 days. Using previously described methods ^9^, we inoculated viral cultures with this spiked milk and examined them for cytopathic effect (normally easily identified by 4 days of culture). After 4 days of culture, RNA was extracted from the culture supernatants and examined using RT-PCR for evidence of SARS-CoV-2 replication.

### Statistical analysis

Maternal and infant characteristics were compared between the group of mothers with confirmed infection for whom no milk samples were positive and those with either confirmed infection, or who were symptomatic and who had at least one milk sample that was positive for viral RNA. Comparisons for continuous variables were made using the Wilcoxon rank-sum test. Categorical variables were compared using Fisher’s exact test. Missing values were excluded. SPSS version 25 was used for analyses, and Prism version 8.4.3 (GraphPad) was used for figure presentation.

## RESULTS

### Detection of SARS-CoV-2 RNA in the breast milk of women

Breast milk samples were available from 110 women: 36 women with SARS-CoV-2 symptoms but no diagnostic test, 9 women with symptoms but a negative nasal or nasopharyngeal SARS-CoV-2 RT-PCR test and 65 women with confirmed SARS-CoV-2 using a nasal or nasopharyngeal RT-PCR test. A total of 336 breast milk samples were collected. Participating women submitted a median of 2.5 samples of breast milk (range 1 to 13). Out of 336 samples, 285 breast milk samples (85%) were available and analyzed for SARS-CoV-2 viral RNA. A total of 160 samples were cultured for virus: 118 from 50 women with confirmed SARS-CoV-2 infection and 42 from the two groups of symptomatic women. All subsequent descriptors presented below are restricted to the 65 women who had a confirmed diagnosis of SARS-CoV-2 and 1 asymptomatic woman whose milk sample was positive for viral RNA, but who was not tested for SARS-CoV-2. We will refer to this group as the 66 women with confirmed SARS-CoV-2 infection.

The majority of women were non-Hispanic and white with a median age of 35.8 years (Table 1). Among these, 59 (89%) had symptomatic COVID-19. Only 6 (9.1%) were hospitalized, and of these, two developed respiratory failure and required ECMO support. Two-thirds (66.7%) of the women hospitalized were pregnant at the time of their SARS-CoV-2 infection and hospitalizations. The two women with severe disease requiring ECMO ultimately delivered their infants while receiving treatment.

**Table 1.** Characteristics of 66 Women with Confirmed SARS-CoV-2 infection^a^

|  | No Detectable SARS-CoV-2<br>in Breast Milk Samples<br>(N = 59) | Detectable SARS-CoV-2<br>in Breast Milk Samples<br>(N = 7) | P-value |
| --- | --- | --- | --- |
| Maternal Age (years) | 35.80 [25.56, 46.67] | 32.28 [27.62, 38.66] | 0.120 |
| Race |  |  | 1.000 |
| Caucasian | 48 (84.21) | 7 (100.00) |  |
| Black | 1 (1.75) | 0 (0.00) |  |
| Asian | 5 (8.77) | 0 (0.00) |  |
| Native American/Alaska Native | 2 (3.51) | 0 (0.00) |  |
| Multiracial | 1 (1.75) | 0 (0.00) |  |
| Ethnicity |  |  | 0.528 |
| Hispanic | 5 (8.77) | 1 (14.29) |  |
| Non-Hispanic | 52 (91.23) | 6 (85.71) |  |
| US Region of Residence |  |  | 0.019 |
| North-East | 21 (35.59) | 1 (14.29) |  |
| South | 15 (25.42) | 3 (42.86) |  |
| Mid-West | 6 (10.17) | 3 (42.86) |  |
| West | 17 (28.81) | 0 (0.00) |  |
| BMI $\geq 30$ | 20 (35.08) | 0 (0.00) | 0.096 |
| Underlying Health Condition<sup>b</sup> | 15 (26.32) | 2 (28.57) | 1.000 |
| Symptomatic | 53 (89.83) | 6 (100.00) | 1.000 |
| Hospitalized | 6 (10.71) | 0 (0.00) | 1.000 |
| Treatment Received: Remdesivir | 3 (5.26) | 0 (0.00) | 1.000 |
| Treatment Received: ECMO | 2 (3.51) | 0 (0.00) | 1.000 |
| Source of Infection |  |  | 0.517 |
| Household | 17 (29.31) | 3 (42.86) |  |
| Community | 11 (18.97) | 2 (28.57) |  |
| Occupational | 18 (31.03) | 2 (28.57) |  |
| Unknown | 12 (20.69) | 0 (0.00) |  |
| Infant Age (months) | 7.97 [0.10, 35.28] | 9.63 [2.79, 15.65] | 0.498 |
| Infant Sex |  |  | 0.691 |
| Male | 25 (43.86) | 4 (57.14) |  |
| Female | 32 (56.14) | 3 (42.86) |  |

<sup>a</sup> 65 who tested positive by RT-PCR of NP specimen and 1 without diagnostic testing whose breast milk sample was positive by RT-PCR
<sup>b</sup> Underlying medical conditions included the following: Asthma, Diabetes Type II, Heart Defect, Hypertension, Hypothyroidism, Irritable Bowel Disease, Kidney Defect, Obesity and Tachycardia

SARS-CoV-2 viral RNA was detectable in one milk sample each from 7 of the 66 with confirmed infection (Figure). The women with detectable SARS-CoV-2 RNA in their breast milk did not differ in age, race or ethnicity, or other demographic parameters from women with negative breast milk samples (Table 1). For the seven women whose milk samples contained viral RNA, SARS-CoV-2 RNA was not detected in the next specimen of milk available, ranging from 1 to 97 days later (Table 2, Figure). There was no clinical evidence of infection among any of the infants being breastfed by the seven women with documented SARS-CoV-2 RNA in breast milk.

**Table 2.** Summary of virologic data from breast milk samples with detectable SARS-CoV-2 RNA

| **Participant** | **Number of Symptoms** | **Number of Days Symptomatic** | **Symptomatic at Time of Sample Collection** | **SARS-CoV-2 RNA in Milk Samples (copies/mL)** | **sgRNA** | **Viral Culture** |
| --- | --- | --- | --- | --- | --- | --- |
| 8* | 9 | 18 | Yes | 25,100 | Negative | Negative |
| 22 | 10 | 17 | Yes | 2,400 | Negative | Negative |
| 24 | 16 | 65 | Yes | 3,230 | Negative | Negative |
| 27** | 9 | 13 | Yes | 10,000 | Negative | Negative |
| 42 | 8 | 17 | Yes | 12,000 | Negative | Negative |
| 47 | 5 | 4 | Yes | 11,200 | Negative | Negative |
| 49 | 7 | 7 | Yes | 1,050 | Negative | Negative |

\*Previously reported
\*\*Woman was symptomatic but not tested

### Cultures and RT-RT-PCR reveal no evidence of infectious SARS-CoV-2 in breast milk

None of the viral cultures of 160 breast milk specimens were positive. We used a real-time RT-PCR assay to further analyze the 7 milk specimens found to contain vRNA for the presence of subgenomic RNA (Table 2). Subgenomic RNAs detected by this assay are spliced RNAs that are produced during coronavirus replication ^15^. The milk specimens were clarified by centrifugation and RNA was extracted from the skimmed milk as before. SgRNA was not detected in any of these 7 specimens.

### Stability of SARS-CoV-2 in human milk

We considered the possibility that freezing and thawing of breast milk inactivates SARS-CoV-2 and prevents its detection by culture. We therefore added a small amount (100 TCID_50_) of infectious virus to milk samples from two healthy women, performed serial freeze-thaw cycles, and then stored the samples at 4°C for 3 days. After this prolonged storage, we added the milk to viral cultures. After 4 days, we removed culture supernatants and used RT PCR to test for the presence of viral RNA. Cultures inoculated with samples that had undergone as many as 3 freeze and thaw cycles remained strongly positive. Cytopathic effect was seen in these cultures and positive control wells inoculated with virus alone. These experiments demonstrated that the temperature excursions that frequently occur during handling of breast milk would not prevent us from detecting infectious SARS-CoV-2.

## DISCUSSION

Breast milk is an invaluable source of nutrition to infants ^16,17^ and contains factors that generally prevent infectious disease. However, breast feeding is an acknowledged means by which HIV and HTLV (Human T cell lymphotrophic virus) can be transmitted to infants. In contrast to these pathogens, Hepatitis B virus and Hepatitis C virus infections produce chronic infections in women and yet are not contraindications to breastfeeding ^18^.

A few small case reports have described the detection of SARS-CoV-2 RNA in breast milk^5,19^and one report suggested the possibility of vertical transmission, although contamination could not be ruled out as the source ^20^. Prior reports have generally examined the breast milk of infected mothers for the presence of the virus using RT-PCR methods.^9,20-28^. In one case, SARS-CoV-2 was said to be detectable by RT-PCR in 4 milk samples collected between 6 and 10 days after the mother’s first positive test. The authors indicated there was no evidence that contamination of the milk samples could have been the source of the virus. However, these reports do not give an overall estimate of frequency of viral RNA and infectious SARS-CoV-2 in human milk, and the report of possible transmission did not exclude the possibility of contamination of milk occurring at the time of collection^19^. The rationale for this study was to determine how often SARS-CoV-2 viral RNA was present in breast milk samples and to examine the risk of infection to infants through breast milk. This study is unprecedented in the use of viral cultures to examine a very large number of breast milk specimens, a limitation of prior studies cited by Lackey et al^19^.

We found that SARS-CoV-2 RNA is seldom detected in breast milk samples from women with confirmed SARS-CoV-2 infection. Moreover, our longitudinal follow-up indicates that even when it is detected, it is an unlikely source of infection for the breastfed baby: viral RNA was only transiently present and we were unable to culture SARS-CoV-2 from any sample.

This study had several limitations as well as strengths. The collection of breast milk samples was not directly observed and we relied on the maternal report of SARS-CoV-2 test results, symptoms and treatments received. However, all participants completed a semi-structured interview guided by trained study staff who prompted for specifics with the aid of a calendar. In addition, to our knowledge, this study represents the largest number of breast milk samples analyzed to date from SARS-CoV-2 infected women. We demonstrated that the SARS-CoV-2 maintains its infectivity despite repeated freezing and thawing and storage at 4°C. In addition, while sgRNA (a potential indicator of virus replication) was not detected in any of the milk specimens already known to contain SARS-CoV-2 RNA, this assay is only moderately sensitive: sgRNA is present in only about half of nasopharyngeal specimens with positive viral cultures (^13,15^and unpublished data by PK).

## Conclusion

These data provide substantial evidence that breastfeeding from women proven or suspected to have had SARS-CoV-2 infection does not represent a hazard for infants.

## Data Availability

Data access may be provided, with appropriate ethics approval, by contacting the authors.

## Acknowledgements

The authors gratefully acknowledge the participation of the women and their infants described in this study.

## Abbreviations

SARS-CoV-2: Severe acute respiratory syndrome associated coronavirus type 2
MIS-C: Multisystem inflammatory syndrome in children
TCID_50_: Median Tissue culture infectious dose
Vero-E6 cells: *Cercopithecus aethiops*–derived epithelial kidney cells
sgRNA: subgenomic RNA
ECMO: Extracorporeal membrane oxygenation

---

**Figure.**
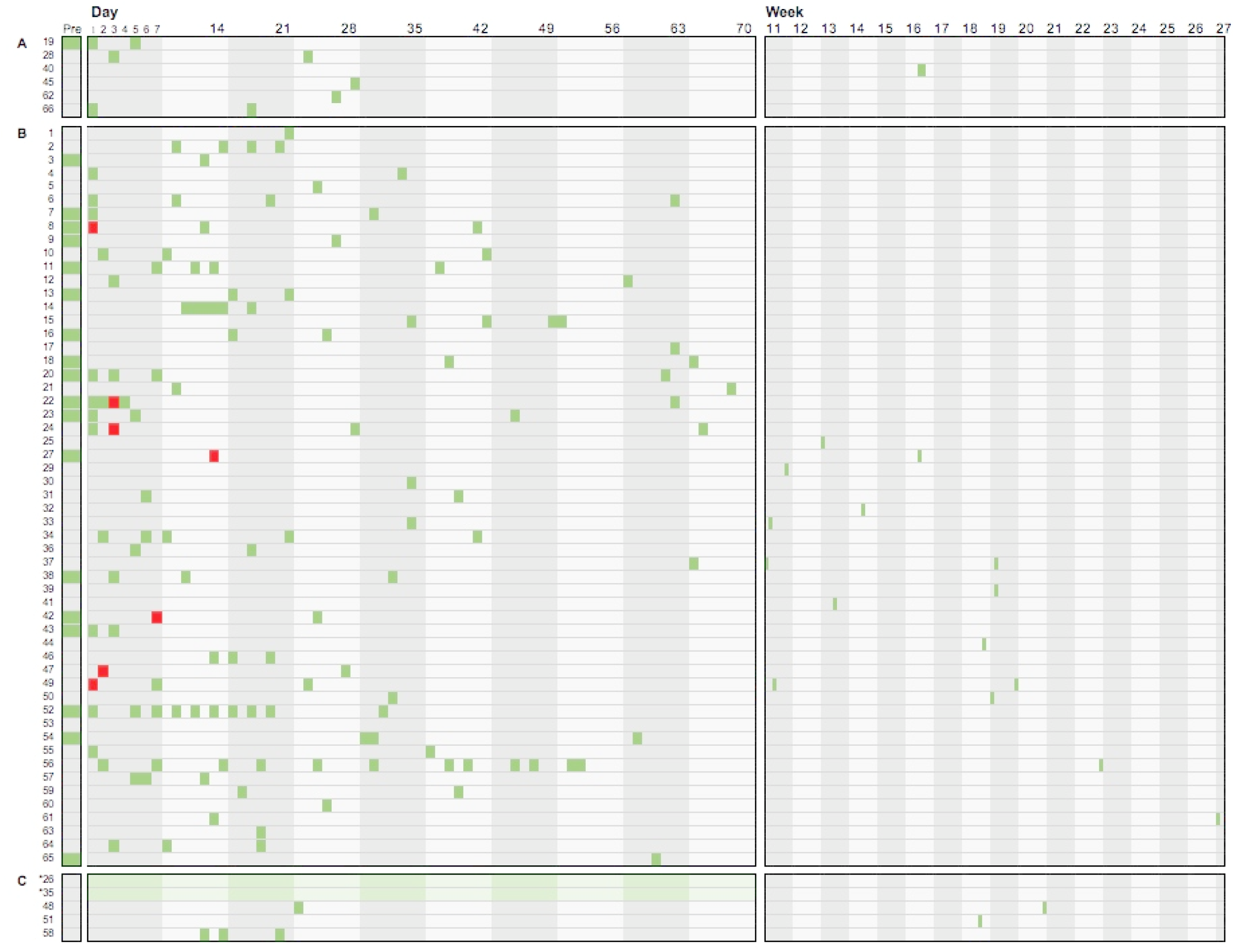
Sample collection timing by day/week from onset of symptoms or hospitalization for 65 women who tested positive for SARS-CoV-2 infection and 1 woman who was symptomatic but untested. Block A displays the timing of breast milk samples collected from women who lacked any symptoms of COVID-19 (SARS-CoV-2 infection) Block B displays the timing of breast milk samples collected by symptomatic women who were not hospitalized. Block C displays are those who were symptomatic and hospitalized. Each colored rectangle demonstrates the time of collection of breast milk specimens in relation to their onset of SARS-CoV-2 symptoms or for those women who were asymptomatic, in relation to their positive test date. Red squares indicate specimens in which the RNA of SARS-CoV-2 was detected by RT-PCR. Green shading indicates women who required ECMO support.

